# Preterm birth rates among twins during the Danish COVID-19 lockdown and mitigation period

**DOI:** 10.1101/2022.09.02.22279489

**Authors:** Paula L Hedley, Marie Bækvad-Hansen, Gitte Hedermann, Henrik Hjalgrim, David M. Hougaard, Mads Melbye, Morten Breindahl, Lone Krebs, Anders Hviid, Ulrik Lausten-Thomsen, Michael Christiansen

**Author notes:** **Correspondence:** Michael Christiansen, MD, FRCPath, Dept for Congenital Disorders, Statens Serum Institut, 5 Artillerivej 2300 S, Copenhagen, Denmark.

## Abstract

Denmark’s response to the COVID-19 pandemic was to issue guidelines on containment, isolation, and surveillance. Subsequently, Denmark entered a period with variable mitigation measures including closures of schools and workplaces, travel restrictions, and restrictions on public gatherings. A Danish study covering the lockdown period (March 12 – April 14, 2020) as well as a period of mitigation measures (February 27 – September 30, 2020) showed that the reduction in extremely preterm births was not associated with an increase in singleton stillbirth rate. Subsequent studies examining the impacts of COVID-19 mitigation measures on preterm birth have combined singleton with multifetal pregnancies. However, multifetal pregnancies have a different set of risk factors for preterm birth, as well as, increased risk of adverse outcomes, and a shorter expected pregnancy length. We assessed the impact of the Danish lockdown period, or the mitigation period on multifetal births and found no significant difference in the proportion of preterm twin births among all twin births, either within gestational age groups or in total, for either period.

---

The Danish response to the COVID-19 pandemic was to issue guidelines on containment, isolation, and surveillance on February 27, 2020, followed by implementation of a lockdown from March 12, 2020 to April 14, 2020. Subsequently, Denmark entered a period with variable mitigation measures including closures of schools and workplaces, travel restrictions, and restrictions on public gatherings, as detailed in ^1^. All restrictions ended March 2022. A Danish nationwide study covering the lockdown period as well as a period of mitigation measures (until end of September 2020) showed that the reduction in extremely preterm births (xPTBs), noted during the strict lockdown period ^2^, was not associated with an increase in singleton stillbirth rate. Rather, between February 27, 2020 and September 31, 2020, the singleton stillbirth rate was ~ 30 % reduced, and the neonatal mortality rate was similar to pre-pandemic levels ^1^. Whereas the finding of reduced xPTBs or preterm births during COVID-19 lockdown was confirmed in several nationwide studies ^1^, however, the reductions were not universal ^1,3^. This may reflect the very different consequences that COVID-19 restrictions had on prenatal care provisions, as well as societal and economic support for pregnant women globally. Likewise, the stringency of COVID-19 mitigation measures, as well as the behavioural, e.g. mobility patterns, and psychological responses to these, varied considerably between countries and over time ^1^. The COVID-19 lockdown constitutes an important quasi-experiment with respect to identification of novel methods to prevent preterm birth (PTB) ^3^.

Three subsequent studies, which included Danish data, all examined PTB rates without excluding multifetal pregnancies ^3-5^. This makes data interpretation difficult, as multifetal pregnancies have a different set of risk factors for PTB, increased risk of adverse outcome, and a shorter expected pregnancy length. This prompted us to examine the effect of the COVID-19 mitigation measures on the PTB rate and gestational age distribution in twin pregnancies.

Using data from the Danish Neonatal Screening Biobank, we calculated the number of the twin births from February 27 – September 30, 2015 – 2020. This enabled us to perform a calendar-matched comparison of the gestational age distribution of twin births, during the strict lockdown, March 12 – April 14, 2020, and the mitigation period, February 27 – September 30, 2020, with the 2015 – 2019 period. The proportion of preterm twin births among all live-born births, who survived to have a blood-spot sample taken 48-72 hrs after birth, was 1.19 % in the lockdown period in 2020, marginally lower than the 1.38 % in 2015-2019. Furthermore, neither the proportion of all live twin births (p = 0.106; ANOVA) nor preterm twin births (p = 0.733, ANOVA), among all live births, exhibited a significant change with year from 2015 to 2019.

There was no significant difference in the proportion of preterm twin births among twin births, either by gestational age groups or in total, during the strict lockdown period from March 12 – April 14, 2020, or the mitigation period from February 27 – September 30, 2020, compared to the 2015 – 2019 period (Table 1). The odds ratio (OR) in xPTBs twins during the strict lockdown period compared to 2015 – 2019 was 0.61 (95% CI, 0.01 – 11.92), and not significant (Table 1). Thus, with the proviso that the confidence intervals are very wide due to the low number of twin births, the COVID-19 lockdown and mitigation measures does not seem to have had a significant effect on the overall preterm birth and xPTB rates of twin pregnancies.

**TABLE 1.** Live twin births in the 2020 lockdown and mitigation periods compared to aggregate data from 2015 – 2019. The aggregate data of twin and singleton births has been included for comparison purposes.

| GA | GA | Live born twin births |  |  | Live born singleton and twin infants<br>(twins counted twice) |  |  |
| --- | --- | --- | --- | --- | --- | --- | --- |
| Category | Weeks + days | 2020 | 2015 - 2019 | 2020 vs 2015-2019 | 2020 | 2015 - 2019 | 2020 vs 2015-2019 |
| **Lockdown (March 12 – April 14)** |  |  |  |  |  |  |  |
|  |  | N (%) | N per year<br>Mean (SD) | OR (95% CI) | N (%) | N per year<br>Mean (SD) | OR (95% CI) |
| Extremely preterm |  | < 5 | < 5 | 0.61 (0.01, 11.92) | < 5 | 15 (3.67) | 0.20 (0.04, 0.72) |
| Very preterm | 22+0 – 31+6 | 6 (9.0) | 6.2 (2.28) | 1.23 (0.31, 4.85) | 44 (0.8) | 42 (5.10) | 1.07 (0.70, 1.64) |
| Moderate preterm | 32+0 – 36+6 | 24 (35.8) | 28.8 (4.87) | 1.02 (0.49, 2.11) | 269 (5.0) | 283 (17.4) | 0.97 (0.82, 1.15) |
| Term | 37+0 – 41+6 | 36 (53.7) | 43.2 (11.9) | 1.03 (0.51, 2.07) | 4917 (91.0) | 4964 (207) | 1.15 (1.02, 1.31) |
| Late term | ≥ 42+0 | < 5 | < 5 | Not estimable | 103 (1.9) | 114 (18.2) | 0.92 (0.71, 1.21) |
| NA |  | < 5 | < 5 |  | 67 (1.2) | 112 (20.0) |  |
| **Period of COVID-19 mitigations (February 27 – September 30)** |  |  |  |  |  |  |  |
|  |  | N (%) | Mean (SD) | OR (95% CI) | N (%) | Mean (SD) | OR (95% CI) |
| Extremely preterm | < 27+6 | 9 (1.7) | 8.2 (1.92) | 1.23 (0.42, 3.70) | 72 (0.2) | 84.6 (6.88) | 0.85 (0.62, 1.16) |
| Very preterm | 28+0 – 31+6 | 35 (6.7) | 35.6 (2.70) | 1.07 (0.64, 1.78) | 262 (0.7) | 265 (11.3) | 0.99 (0.83, 1.17) |
| Moderate preterm | 32+0 – 36+6 | 190 (36.5) | 200 (25.2) | 1.06 (0.82, 1.37) | 1,889 (5.0) | 1,932 (62.2) | 0.97 (0.91, 1.04) |
| Term | 37+0 – 41+6 | 285 (54.7) | 323 (28.8) | 0.92 (0.72, 1.18) | 34,623 (91.0) | 34,080 (988) | 1.14 (1.09, 1.20) |
| Late term | ≥ 42+0 | < 5 | < 5 | Not estimable | 833 (2.2) | 842 (81.0) | 0.99 (0.90, 1.09) |
| NA |  | < 5 | < 5 |  | 368 (1.0) | 732 (108) |  |

GA: gestational age

Studies including twin pregnancies ^3-5^ will tend to underestimate an effect solely acting on singleton pregnancies. The OR is 0.20 (95% CI, 0.04 – 0.72) for xPTB rates of combined singleton and twin pregnancies (Table 1), higher, albeit not significantly, than the reported OR 0.09 (95% CI, 0.01 – 0.40) (2) for singleton births during the lockdown period. As the prenatal care of twin pregnancies, including recommended gestational age of birth, as well as physiological drivers of parturition differ from those characteristic of singleton pregnancies, inclusion of multifetal pregnancies may obscure significant findings among singleton pregnancies. This will be of particular significance for births ≤ 32 weeks (Table 1) and ^1^, where the proportion of twins is ~ 10 %, as opposed to the ~ 1 % of all pregnancies.

As an illustration of the impact of combining multifetal with singleton births, a study based on data from several international Neonatal Intensive Care Units (NICUs) reported a decrease in NICU admissions of xPTBs, including multifetal births, in Denmark and Norway of 18 % during the most restrictive three-month mitigation period ^4^. If it is accepted that Norway did not experience a reduction in xPTBs ^3^, assuming a 10 % proportion of multifetal xPTB pregnancies, then the Danish reduction of singleton xPTBs pregnancies over a three-month period can be estimated to be ~ 40 %. Clearly compatible with the 73 % (95% CI, 14 % - 93 %) reduction in xPTBs found in the Danish National Patient Register based study on all Danish singleton preterm births ^1^, and the 56 % and 47 % reductions in xPTBs in March and April 2020 reported in a study based on data from the Danish Newborn Quality Database ^5^.

In conclusion, the COVID-19 quasi-experiment did not seem to influence the gestational age distribution of multifetal pregnancies. In Denmark, this may reflect that women with a multifetal pregnancy are subject to an intensive prenatal care regimen that increases resilience to stress and anxiety. This is speculative, and no causal relations have been identified; however, an association between maternal anxiety and preterm birth has been reported repeatedly over the last four decades ^1^. Furthermore, it seems that individual phenotyping, e.g. clarification of individual risk factors, could be a way forward to identify pregnancies where interventions may reduce the risk of PTB. Finally, the consequences of combining singleton and multifetal pregnancies should always be considered, particularly when studying adverse outcomes known to occur frequently in multifetal pregnancies.

## Data Availability

All data produced in the present work are contained in the manuscript

## ETHICS

The study was conducted according to Danish legislation and guidelines for register research and was approved by the Data Protection Agency officer at Statens Serum Institut (No: 20/04753).

## ACKNOWLEDGEMENTS

This research was conducted using data obtained from the Danish National registers. The Danish Health Authority is acknowledged for support with accessing data.

